# Accumulating Sedentary Time and Physical Activity from Childhood to Adolescence and Cardiac Function in Adolescence

**DOI:** 10.1101/2023.07.19.23292912

**Authors:** Eero A. Haapala, Marja H. Leppänen, Earric Lee, Kai Savonen, Jari A. Laukkanen, Mika Kähönen, Soren Brage, Timo A. Lakka

## Abstract

**Background:** Little is known about the associations of a sedentary lifestyle from childhood with cardiac work and function during adolescence. We studied if cumulative sedentary time and physical activity from childhood to adolescence are associated with cardiac work and function in adolescence.

**Methods:** A total of 153 adolescents aged 15 years at the time of assessment of cardiac work and function participated. We assessed sedentary time and physical activity using a combined movement and heart rate sensor in childhood and adolescence (baseline, 2- and 8-year follow-ups) and cardiac work and function using impedance cardiography in adolescence (8-year follow-up).

**Results:** Cumulative sedentary time from baseline over the follow-up was directly (standardised regression coefficient β=0.245 to 0.246, 95% confidence intervals, CI=0.092 to 0.400) and moderate to vigorous physical activity (β=-0.355 to -0.323, 95% CI=-0.579 to - 0.119), and vigorous physical activity (β=-0.305 to -0.295, 95% CI=-0.524 to -0.083) from baseline over the follow-up was inversely associated with cardiac work at 8-year follow-up. Cumulative vigorous physical activity from baseline to 2- and 8-year follow-up was inversely associated with cardiac work index at 8-year follow-up (β=-0.225 to -0.218, 95% CI=-0.450 to 0.000). However, adiposity and other cardiometabolic risk factors partially explained these associations.

**Conclusions:** Higher levels of sedentary time and lower levels of moderate and/or vigorous-intensity physical activity during childhood were associated with higher cardiac work in adolescence. These findings highlight the importance of obesity prevention and weight management and promotion physically active lifestyle since childhood to prevent abnormalities in cardiac function later in life.

**CLINICAL PERSPECTIVE:**

- Sedentary lifestyle increases the risk of cardiovascular diseases, but little is known about the role of sedentary time and physical activity in cardiac work and function in youth.
- We found that adolescents accumulating higher levels of sedentary time and lower levels of physical activity since childhood had higher cardiac workload compared to their more physically active peers. However, these associations were partly explained by adiposity and other cardiometabolic risk factors.
- These findings highlight the importance of obesity prevention and weight management and promotion physically active lifestyle since childhood to prevent abnormalities in cardiac function later in life.

## INTRODUCTION

Adolescents are growing increasingly sedentary (1), and less than 20% of them achieve the recommended levels of moderate to vigorous physical activity (MVPA) (2). Increased time spent in sedentary behaviours, i.e., in activities with minimal movement and low energy expenditure (3), may increase the risk of atherosclerotic cardiovascular diseases (4). Conversely, increased PA has been suggested to counteract the undesirable effects of sedentary behaviour on cardiovascular health (5). However, whether a sedentary lifestyle since childhood is associated with increased cardiac work and impaired cardiac function in adolescence, increasing the risk of cardiac insufficiency later in life, is poorly understood. Although studies among adults suggest that prolonged bed-rest negatively alters cardiac structure and function (6, 7), observational studies have reported weak associations between sedentary time and cardiac structure or function (8–10). In contrast, higher levels of MVPA and vigorous PA have been associated with better cardiac functions in adults (10, 11). Additionally, high and moderate-intensity interval training have been associated with improved resting cardiac function (12, 13). Regular PA has also been associated with reduced myocardial workload (13).

The development of cardiovascular diseases and early cardiac damage requires a relatively long exposure to an unhealthy lifestyle, often beginning in childhood (Steinberger *et al.*, 2016). Moreover, cardiac morphology and function continuously develop and change during growth (15). Although mixed findings have been reported on the associations between sedentary time and arterial health in children and adolescents (16), the role of sedentary time in cardiac work and function has received less attention (17, 18). Furthermore, the association between PA from childhood with cardiac work and function in adolescence has yet to be explored.

Low circulating HDL cholesterol levels have been associated with an impaired cardiac function (19). Higher serum HDL may contribute to cardiac functions by reducing stress-induced myocardial hypertrophy, cell injury, and regulating intracellular signalling pathways (20). Moreover, sedentary lifestyle and physical inactivity have been found to induce insulin resistance, increase blood pressure, and arterial stiffness, potentially altering cardiac work and function. However, their role in explaining the associations of sedentary time and PA with cardiac work and function remains unknown (13, 21, 22).

Most paediatric studies on the associations of sedentary time and PA with cardiovascular health have been cross-sectional or included only a short follow-up period and have focused on arterial structure and function (16, 18). Therefore, we studied the associations of cumulative sedentary time and PA from childhood to adolescence with cardiac work and function in adolescence over an 8-year follow-up period. We also investigated whether body fat percentage (BF%), insulin resistance, systolic blood pressure (SBP), arterial stiffness, or HDL characteristics modify the observed associations.

## METHODS

### Study design and participants

The present data are from the Physical Activity and Nutrition in Children (PANIC) study, which is an 8-year physical activity and dietary intervention study and a long-term follow-up study in a population sample of children from the city of Kuopio, Finland (23). The Research Ethics Committee of the Hospital District of Northern Savo approved the study protocol in 2006 (Statement 69/2006). The parents or caregivers of the children gave their written informed consent, and the children provided their assent to participation. The PANIC study has been carried out in accordance with the principles of the Declaration of Helsinki as revised in 2008.

Altogether 736 children aged 6–8 years from primary schools of Kuopio were invited to participate in the baseline examination in 2007–2009. A total of 512 children, who represented 70% of those invited, participated in the baseline examinations. Six children were excluded from the study at baseline because of physical disabilities that could hamper participation in the intervention or had no time or motivation to attend the study. The participants did not differ in sex distribution, age, or body mass index standard deviation score (BMI-SDS) from all children who started the first grade in 2007–2009 based on data from the standard school health examinations performed for all Finnish children before the first grade. We conducted the main analyses for 153 participants (63 girls, 84 boys) who had valid data on cardiac work and function at 8-year follow-up and valid sedentary time and PA data at least at one time point.

### Assessment of cardiac work and function

Cardiac work (kg x m), stroke volume (mL), and cardiac output (L/min) were measured with the Circmon^®^ B202 impedance cardiography device (JR Medical Ltd, Saku Vald, Estonia) after a 15 minute supine rest (24, 25). These measures were also normalised for body surface area and expressed as cardiac word index (kg x m/m^2^), stroke index (mL/m^2^), and cardiac index (L/min/m^2^). The method and electrode configuration have been described in detail elsewhere (24). Cardiac work index was calculated using the formula: 0.0143 x (mean aortic pressure–pulmonary artery occlusion pressure) x cardiac index. Pulmonary artery occlusion pressure was assumed to be 6 mmHg, and 0.0143 is the conversion factor of pressure from millimetres of mercury to centimetres of water, volume to density of blood (in kg/L), and centimetres to metres (25). The stroke volume and cardiac output measured using Circmon whole-body impedance cardiography agree reasonably well with values measured using 3-dimensional ultrasound and the thermodilution method (24, 26, 27).

### Assessment of sedentary time and physical activity

Sedentary time and PA were assessed using a combined heart rate and movement sensor (Actiheart®, CamNtech Ltd., Papworth, UK) for a minimum of four consecutive days without interruption, including sleep and water-based activities, analysed in 60 s epochs, as described previously (28, 29). We defined the valid data as a recording period of at least 48 h of wear data with the additional requirement that enough data were included from all four quadrants of a 24 h day to avoid bias from over-representation of specific parts of days (30). This resulted in at least 12 h of wear data from morning (3 am – 9 am), noon (9 am – 3 pm), afternoon / evening (3 pm – 9 pm), and night (9 pm – 3 am).

Data on heart rate were cleaned and individually calibrated with parameters from the maximal exercise test and combined with movement sensor data to derive PA energy expenditure (PAEE). Instantaneous PAEE, i.e. PA intensity, was estimated using branched equation modelling as explained, in detail earlier (31) and summarised as daily PA volume (kJ·day^−1^·/kg^−1^) and time spent at certain levels of standard metabolic equivalents of task (METs) in minutes per day, weighting all hours of the day equally to reduce diurnal bias caused by imbalances in wear-time. Initially, the summarised data included 25 narrowly defined intensity categories. For the present analyses, we re-categorised these intensity categories into a broader format of sedentary time (≤1.5 METs), light PA (1.5-4 METs), moderate PA (>4-7 METs), MVPA (>4 METs), and vigorous PA (>7 METs), which have been commonly applied in investigations of PA among children and youth. Combined heart rate and movement sensing has been found to be more accurate in estimating PAEE than either method alone in children (32, 33), explaining 86% of variance in PA energy expenditure variance (33).

We used the AUC for sedentary time and PA measured at baseline, 2-year follow-up, and 8-year follow-up to utilise all data collected over the 8-year period and to describe the exposure to sedentary time and PA from childhood to adolescence (34). The AUCs were determined using an additive mixed model (35). The modelling allowed the inclusion of a non-linear effect which was modelled by cubic spline in addition to random intercept for individuals (36). For this study, the AUC variables for sedentary time and PA were defined separately for childhood (from baseline to 2-year follow-up) and from childhood to adolescence (from baseline to 8-year follow-up).

Because the Area Under the Curve (AUC) approach used to quantify cumulative sedentary time and PA uses estimated data, we performed the sensitivity analyses among 81 participants (28 girls, 53 boys) who had valid and complete data on sedentary time and PA at all three time points. We performed additional sensitivity analyses using mean sedentary time and PA from baseline to 2-year follow-up and from baseline to 8-year follow-up among 81 participants (28 girls, 53 boys) who had valid and complete data for sedentary time and PA in all time points. This was performed to account for the reduction in the reliability of the data due to the long interval from the assessment of sedentary time and PA from childhood to adolescence (37).

### Assessment of modifying factors

Whole body mass was measured twice with the children having fasted for 12 hours, emptied the bladder, and standing in light underwear by a calibrated InBody^®^ 720 bioelectrical impedance device (Biospace, Seoul, South Korea) to an accuracy of 0.1 kg. The mean of these two values was used in the analyses. Stature was measured three times with the children standing in the Frankfurt plane without shoes using a wall-mounted stadiometer to an accuracy of 0.1 cm. The mean of the nearest two values was used in the analyses. BMI was calculated by dividing body mass (kg) by body height (m) squared. BMI-SDS was calculated based on Finnish reference data (38). The prevalence of overweight and obesity was defined using the cut-off values provided by Cole et al. (39). Total fat mass, BF%, and lean mass (LM) were measured by the Lunar^®^ dual-energy X-ray absorptiometry device (GE Medical Systems, Madison, WI, USA) using standardised protocols (40).

SBP was measured from the right arm using the Heine Gamma G7^®^ aneroid sphygmomanometer (Heine Optotechnik) to an accuracy of 2 mm Hg. The measurement protocol included a 5 min seated resting period followed by three measurements with 2 min intervals in between. The average of all three values was used in the analyses. Pulse wave velocity was assessed by the Circmon^®^ impedance cardiography.

A research nurse took blood samples in the morning, after children had fasted overnight for at least 12 hours. Blood was immediately centrifuged and stored at a temperature of -75°C until biochemical analyses. Plasma glucose was measured by a hexokinase method and serum insulin was measured by an electrochemiluminescence immunoassay. Intra-assay and inter-assay coefficient of variation for the insulin analyses were 1.3–3.5% and 1.6–4.4%, respectively. Insulin resistance was assessed using Homeostatic Model Assessment for Insulin Resistance (HOMA-IR) and the formula fasting serum insulin x fasting plasma glucose/22.5) (41). The Nightingale high-throughput throughput nuclear magnetic resonance spectroscopy platform was used to assess HDL cholesterol, average HDL diameter, and the concentrations of extra-large, large, medium, and small HDL particles, and the concentration of Apolipoprotein A1 (42).

A research physician assessed pubertal status using a 5-stage scale described by Marshall and Tanner (43, 44). We used testicular volume assessed by an orchidometer to assess pubertal status in boys and breast development to assess pubertal status in girls.

### Statistical methods

Statistical analyses were performed using the SPSS statistical software, version 28.0 (IBM corp. Armonk, NY, USA). The characteristics of children between boys and girls were compared using the Student’s t-test for normally distributed continuous variables, the Mann-Whitney’s U-test for continuous variables with skewed distributions, or the χ^2^-test for categorical variables. Before the analyses, we performed square root or natural logarithm transformation for skewed variables. The associations of cumulative sedentary time and PA with the measures of cardiac work and function were investigated using linear regression analyses adjusted for age and sex. These data were further adjusted for possible modifiers of the associations including pubertal status, BF%, HOMA-IR, SBP, arterial stiffness, or HDL characteristics, which were entered to models separately. We replaced missing data on these measures using the sample mean. To study the modifying effect of sex on the associations of cumulative sedentary time and PA with measures of cardiac work and function, we used general linear models including a sex x sedentary time or PA interaction term in the models.

## RESULTS

### Characteristics of participants

Girls had more advanced pubertal development and were shorter and lighter compared to boys (Table 1.). Girls also had higher BF%, lower SBP, and higher plasma levels of HDL cholesterol, total, extra-large, large, and medium HDL particles, and Apolipoprotein A1 than boys. Girls accumulated less MVPA and vigorous PA than boys.

**Table 1.** Characteristics of participants at 8-year follow-up.

|  | All | Girls | Boys | P-value |
| --- | --- | --- | --- | --- |
| Age (years) | 15.8 (15.5–16.1) | 15.8 (15.4–16.1) | 15.8 (15.5–16.2) | 0.328 |
| Height (cm) | 171.9 (8.1) | 166.6 (5.4) | 176.3 (7.2) | **<0.001** |
| Weight (kg) | 59.5 (54.6–67.0) | 57.2 (52.7–63.1) | 63.3 (56.2–73.2) | **<0.001** |
| BMI | 20.6 (18.8–22.0) | 20.9 (19.4–21.9) | 20.1 (18.5–22.1) | 0.188 |
| BMI-SDS | 0.03 (0.9) | 0.16 (0.9) | -0.08 (1.0) | 0.110 |
| Prevalence of overweight/obesity (%) | 15.7 | 14.5 | 16.7 | 0.713 |
| Pubertal status (%) |  |  |  |  |
| 3 | 7.5 | 1.5 | 12.7 | **0.005** |
| 4 | 56.8 | 52.2 | 60.8 |  |
| 5 | 35.6 | 46.3 | 26.6 |  |
| Body fat percentage (%) | 24.2 (13.2–30.6) | 29.5 (24.6–34.3) | 14.6 (10.7–22.8) | **<0.001** |
| HOMA-IR | 2.2 (1.7–3.1) | 2.3 (1.8–3.1) | 2.1 (1.6–3.3) | 0.506 |
| Systolic blood pressure (mmHg) | 113 (11) | 110 (8) | 116 (12) | **<0.001** |
| Pulse wave velocity (m/s) | 5.7 (5.5–6.1) | 5.7 (5.5–6.1) | 5.7 (5.4–6.0) | 0.369 |
| HDL cholesterol (mmol/L) | 1.4 (0.2) | 1.5 (0.2) | 1.2 (0.2) | **<0.001** |
| Concentration of HDL particles (mmol/L) | 0.015 (0.001) | 0.016 (0.002) | 0.014 (0.001) | **<0.001** |
| Average HDL diameter (nm) | 9.7 (0.2) |  |  |  |
| Concentration of extra-large HDL particles (mmol/L) | 0.0002 (0.0002–0.0003) | 0.0002 (0.0002–0.0003) | 0.0002 (0.0001–0.0002) | **<0.001** |
| Concentration of large HDL particles (mmol/L) | 0.001 (0.001–0.002) | 0.002 (0.001–0.002) | 0.001 (0.001–0.002) | **<0.001** |
| Concentration of medium HDL particles (mmol/L) | 0.004 (0.001) | 0.004 (0.001) | 0.003 (0.001) | **<0.001** |
| Concentration of small HDL particles (mmol/L) | 0.009 (0.001) | 0.010 (0.001) | 0.009 (0.001) | 0.070 |
| Apolipoprotein A1 (g/L) | 1.4 (0.18) | 1.5 (0.18) | 1.3 (0.13) | **<0.001** |
| Sedentary time (min/d) | 604 (139) | 609 (142) | 601 (139) | 0.788 |
| Light physical activity (min/d) | 321 (117) | 321 (124) | 322 (113) | 0.970 |
| Moderate to vigorous physical activity (min/d) | 39 (20–68) | 25 (19–45) | 51 (26–71) | **0.009** |
| Vigorous physical activity (min/d) | 5 (1–19) | 2 (0–6) | 9 (1–26) | **0.002** |
| Physical activity energy expenditure<br>(kJ/kg/d) | 55.9 (22.5) | 51.9 (19.6) | 59.1 (24.3) | 0.051 |
| Cardiac work (kg x m) | 5.1 (1.3) | 4.9 (1.1) | 5.2 (1.4) | 0.164 |
| Cardiac work index (kg x m/m<sup>2</sup>) | 2.9 (0.7) | 3.0 (0.7) | 2.9 (0.7) | 0.449 |
| Stroke volume index (mL/m<sup>2</sup>) | 42.1 (6.3) | 42.2 (6.1) | 42.1 (6.6) | 0.930 |
| Cardiac index (L/min/ m<sup>2</sup>) | 2.9 (0.5) | 3.0 (0.5) | 2.9 (0.5) | 0.111 |

The data are means and standard deviations or medians and interquartile ranges\*. P-value for the differences between girls and boys are from Student's t -test, Mann-Whitney U -test, or $\chi^2$ -test. BMI, body mass index; BMI-SDS, body mass index standard deviation score; HDL, high-density lipoprotein; HOMA-IR, Homeostatic Model Assessment for Insulin Resistance.

### Associations of cumulative sedentary time with cardiac work and function

A cumulative sedentary time from baseline to 2-year follow-up and from baseline to 8-year follow-up was directly associated with cardiac work at 8-year follow-up after adjustment for age and sex (Table 2). The associations of cumulative sedentary time from baseline to 2-year follow-up (β=0.154, 95% CI=-0.005 to 0.312) and from baseline to 8-year follow-up (β=0.153, 95% CI=-0.005 to 0.310) with cardiac work attenuated after further adjustment for BF%.

**Table 2.** Associations of cumulative sedentary time and physical activity with cardiac work and function.

|  | Left cardiac work (kg x m) | Left cardiac work index (kg x m/m<sup>2</sup>) | Cardiac index (mL/min/m<sup>2</sup>) | Stroke volume index (mL/m<sup>2</sup>) |
| --- | --- | --- | --- | --- |
| Sedentary time |  |  |  |  |
| 0–2 | **0.246 (0.092 to 0.400)** | 0.158 (-0.001 to 0.317) | -0.003 (-0.164 to 0.158) | 0.143 (-0.016 to 0.302) |
| 0–8 | **0.245 (0.092 to 0.398)** | 0.158 (-0.001 to 0.316) | -0.003 (-0.164 to 0.158) | 0.142 (-0.016 to 0.301) |
| Light physical activity |  |  |  |  |
| 0–2 | -0.192 (-0.385 to 0.001) | -0.123 (-0.319 to 0.074) | 0.066 (-0.132 to 0.263) | -0.094 (-0.291 to 0.103) |
| 0–8 | -0.174 (-0.348 to 0.001) | -0.111 (-0.289 to 0.067) | 0.059 (-0.120 to 0.239) | -0.085 (-0.263 to 0.093) |
| Moderate-to-vigorous physical activity |  |  |  |  |
| 0–2 | **-0.355 (-0.579 to -0.131)** | -0.226 (-0.458 to 0.005) | -0.070 (-0.304 to 0.165) | **-0.251 (-0.481 to -0.021)** |
| 0–8 | **-0.323 (-0.527 to -0.119)** | -0.206 (-0.417 to 0.005) | -0.063 (-0.277 to 0.150) | **-0.229 (-0.439 to -0.019)** |
| Vigorous physical activity |  |  |  |  |
| 0–2 | **-0.305 (-0.524 to -0.085)** | **-0.225 (-0.450 to 0.000)** | 0.043 (-0.185 to 0.271) | -0.164 (-0.389 to 0.062) |
| 0–8 | **-0.295 (-0.508 to -0.083)** | **-0.218 (-0.436 to 0.000)** | 0.042 (-0.179 to 0.263) | -0.159 (-0.377 to 0.060) |
| Physical activity energy expenditure |  |  |  |  |
| 0–2 | **-0.300 (-0.487 to -0.113)** | -0.149 (-0.343 to 0.045) | -0.067 (-0.263 to 0.129) | **-0.209 (-0.401 to -0.017)** |
| 0–8 | **-0.290 (-0.471 to -0.110)** | -0.144 (-0.332 to 0.044) | -0.065 (-0.254 to 0.124) | **-0.202 (-0.388 to -0.016)** |

The data are standardised regression coefficients and their 95% confidence intervals adjusted for age and sex. Statistically significant associations are bolded. 0–2 and 0–8 describe time points used to calculate cumulative exposures.

Higher cumulative sedentary time from baseline to 2-year follow-up and from baseline to 8-year follow-up were associated with lower stroke volume index in girls (β=-0.257, 95% CI=-0.496 to -0.018) but not in boys (β=0.178, 95% CI=-0.038 to 0.394, p=0.009 for interaction). Further adjustment for PWV attenuated the inverse association between sedentary time and stroke volume index in girls (β=-0.213, 95% CI=-0.458 to 0.031). In these analyses, the standardised regression coefficients were identical for both time points.

### Associations of cumulative physical activity with cardiac work and function

Cumulative MVPA and PAEE from baseline to 2-year follow-up and from baseline to 8-year follow-up were inversely associated with cardiac work and cardiac index after adjustment for age and sex (Table 2). Cumulative vigorous PA from baseline to 2-year follow-up and from baseline to 8-year follow-up was inversely associated with cardiac work and cardiac work index.

Further adjustment for BF% attenuated the associations of cumulative MVPA (β=-0.205, 95% CI=-0.439 to 0.029 and β=-0.187, 95% CI=-0.400 to 0.027 for cumulative exposures from baseline to 2-year-follow-up and from baseline to 8-year follow-up, respectively), cumulative vigorous PA (β=-0.171, 95% CI=-0.385 to 0.054 and β=-0.165, 95% CI=-0.382 to 0.468), and cumulative PAEE (β=-0.163, 95% CI=-0.363 to 0.037 and β=-0.158, 95% CI=-0.351 to 0.035) with cardiac work and the association between cumulative vigorous PA and cardiac work index (β=-0.203, 95% CI=-0.443 to 0.036 and β=-0.197, 95% CI=-0.429 to 0.035).

The associations of cumulative vigorous PA with cardiac work index also attenuated after further adjustment for HOMA-IR (β=-0.225, 95% CI=-0.455 to 0.005 and β=-0.218, 95% CI=-0.441 to 0.005), PWV (β=-0.190, 95% CI=-0.141 to 0.033 and β=-0.184, 95% CI=-0.401 to 0.032), small HDL particles (β=-0.224, 95% CI=-0.449 to 0.001 and β=-0.217, 95% CI=-0.435 to 0.001), and APO A1 (β=-0.224, 95% CI=-0.449 to 0.001 and β=-0.217, 95% CI=-0.435 to 0.001).

The associations of cumulative MVPA (β=-0.212, 95% CI=-0.443 to 0.020 and β=-0.193, 95% CI=-0.404 to 0.018) and cumulative PAEE (β=-0.178, 95% CI=-0.371 to 0.015 and β=-0.172, 95% CI=-0.358 to 0.015) with cardiac index attenuated after further adjustment for PWV.

Cumulative light PA (β=0.257, 95% CI=0.020 to 0.494 vs. β=-0.120, 95% CI=-0.338 to 0.097, p=0.026 for interaction) and cumulative vigorous PA (β=0.237, 95% CI=0.000 to 0.474 vs. β=-0.048, 95% CI=-0.367 to 0.171, p=0.059 for interaction) from baseline to 2-year follow-up and from baseline to 8-year follow-up were directly associated with stroke volume index in girls but not in boys. In these analyses, the standardised regression coefficients were identical for both time points.

### Sensitivity analyses

A cumulative sedentary time from baseline to 2-year follow-up and from baseline to 8-year follow-up was directly associated with cardiac work, cardiac work index, and cardiac index at 8-year follow-up (Supplementary Table S1.) A cumulative MVPA from baseline to 2-year follow-up and from baseline to 8-year follow-up was inversely associated with cardiac work. A higher PAEE from baseline to 2-year follow-up and from baseline to 8-year follow-up was associated with lower cardiac work, cardiac work index, and cardiac index.

A higher mean sedentary time from baseline to 8-year follow-up was associated with higher cardiac work, cardiac work index, and cardiac index (Supplementary Table S2.). A higher mean MVPA from baseline to 8-year follow-up was inversely associated with cardiac work. Average PAEE from baseline to 2-year follow-up and from baseline to 8-year follow-up was associated with lower cardiac work. A higher average PAEE from baseline to 8-year follow-up was associated with lower cardiac work index and cardiac index.

Further adjustment for BF%, HOMA-IR, and PWV attenuated the association between MVPA and cardiac work. Further adjustment for SBP and PWV attenuated the associations of sedentary time or PAEE with cardiac work index, respectively.

## DISCUSSION

We observed that higher cumulative sedentary time and lower cumulative MVPA, vigorous PA, and PAEE from childhood to adolescence were associated with higher cardiac work in adolescence. However, only cumulative vigorous PA was inversely associated with the cardiac work index. Nevertheless, these associations between sedentary time, PA, and cardiac work attenuated after controlling for BF%. Interestingly, when we applied tighter inclusion criteria for the valid data for sedentary time and PA in the sensitivity analyses, lower levels of sedentary time and higher PAEE were associated with cardiac work index independent of BF% but not SBP or PWV, respectively. Therefore, our findings suggest that higher cumulative sedentary time and lower cumulative PA from childhood to adolescence are associated with higher cardiac work in adolescence. However, these associations are partly due to variations in cardiometabolic risk factors.

Our findings suggest that increased sedentary time may increase cardiac work and cardiac oxygen demand. Higher levels of sedentary time have been associated with higher left ventricular mass in adolescents (18) and adults (10), indicating possible pathophysiological alterations underlying left ventricular hypertrophy and higher cardiac oxygen consumption Thus, enlargement in cardiac muscle and left ventricular size may explain the present findings of a direct association between sedentary time and cardiac work. Nevertheless, BF% and other cardiometabolic risk factors partly explain the observed associations between sedentary time and cardiac work. Accordingly, obesity increases the cardiac work in youth (45) Therefore, increased adiposity since childhood may have bidirectional associations with sedentary time. As such, the independent role of sedentary time in the alterations of cardiac work and function remains unknown (10). Notwithstanding, in adults with obesity, weight loss has been associated with reduced left ventricular mass (47). Therefore, preventing increased adiposity, insulin resistance, and arterial stiffness by reducing sedentary time and increasing PA may reduce cardiac workload in youth.

In line with previous studies in adults (13), we found that higher levels of PA were associated with lower cardiac work. Moreover, our current observations suggest that at least moderate-intensity PA, but not light-intensity PA, was inversely associated with cardiac work. It is likely that an intensity threshold for PA needs to be met to elicit beneficial cardiac adaptations, which is the same for adults (48). These findings align with observations that higher-intensity PA is more effective in improving cardiovascular functions, such as maximal oxygen uptake (49) and endothelial function in youth (50). Nevertheless, we observed an inverse association between PAEE and cardiac work, suggesting that also the total volume of PA with sufficient intensity is also an important determinant of cardiac work. Contrary to our findings suggesting lower cardiac oxygen consumption, MVPA has been associated with a higher left ventricular mass in adolescents (18). However, the direct associations between MVPA and left ventricular mass may be due to a positive adaptation through athleteś heart as endurance training increases left ventricular mass in youth (51) and adults (52). These positive cardiac adaptations reduce left ventricular wall stress and decrease the cardiac oxygen demand (53), which is in agreement with our findings.

Our findings, in line with the results of some previous studies (53, 54), suggest that sedentary time and PA have weak, if any, associations with resting stroke volume and cardiac output. However, we observed that sedentary time was directly and PAEE was inversely associated with cardiac index. While these results could be related to alterations in cardiac structures and cardiovascular regulation (10, 18, 53), differences in resting heart rate are the most plausible explanation for these observations.

The strengths of the present study include an 8-year follow-up among a relatively large sample of youth using valid and reproducible methods to assess sedentary time, PA, and cardiac work and function. We also controlled several possible confounding factors, allowing us to investigate their role in the associations of sedentary time and PA with cardiac work and function. It would have been optimal to use cardiac magnetic resonance imaging or echocardiography, which are considered the gold standards for assessing cardiac structures and functions. However, stroke volume and cardiac output assessed using impedance cardiography show a reasonable level of agreement with other methods (24, 26, 27). Easily available cardiac work assessment using impedance cardiography could also have clinical value as increased cardiac workload may increase the risk of heart failure (45). We assessed cardiac work and functions only at the 8-year follow-up and not throughout childhood to adolescence. As such, we took a conservative approach both statistically and, in our interpretations, as causality cannot be ascertained. Our sample included apparently healthy Caucasian adolescents and therefore out findings may not be directly generalisable to other populations. Finally, we assessed resting cardiac work and function, and more studies investigating the associations of sedentary time and PA with cardiac work and function during exercise and other stress conditions are warranted.

In conclusion, higher cumulative sedentary time, lower cumulative MVPA, and cumulative vigorous PA from middle childhood to adolescence were associated with higher cardiac work in adolescence. However, adiposity and other cardiometabolic risk factors partially explains these associations. Our results highlight the importance of increasing PA and obesity prevention from childhood to decrease the risk of cardiac-related abnormalities later in life.

Long-term observational and intervention studies are required to establish the role of PA increase and sedentary time reduction in cardiac functions in children and adolescents.

## SOURCES OF FUNDING

The PANIC study has been supported financially by grants from the Research Council of Finland, Ministry of Education and Culture of Finland, Ministry of Social Affairs and Health of Finland, Research Committee of the Kuopio University Hospital Catchment Area (State Research Funding), Finnish Innovation Fund Sitra, Social Insurance Institution of Finland, Finnish Cultural Foundation, Foundation for Paediatric Research, Diabetes Research Foundation in Finland, Finnish Foundation for Cardiovascular Research, Juho Vainio Foundation, Paavo Nurmi Foundation, Yrjö Jahnsson Foundation, and the city of Kuopio.

## DISCLOSURES

The authors declare no conflict of interest.

## AUTHORS’ CONTRIBUTIONS

EAH, conceptualisation, formal analysis, writing original draft, funding acquisition; MHL, conceptualisation, writing review and editing, EL, writing review and editing; KS, methodology, investigation, writing review and editing; JAL, methodology, investigation, writing review and editing; MK, methodology, writing review and editing; SB, methodology, writing review and editing; TAL, supervision, funding acquisition, project administration, methodology, investigation, writing review and editing. All authors approved the final version of the manuscript.

## DATA AVAILABILITY

The data are not publicly available due to research ethical reasons and because the owner of the data is the University of Eastern Finland and not the research group. However, the corresponding author can provide further information on the PANIC study and the PANIC data on a reasonable request.

## SUPPLEMENTAL MATERIAL

Supplemental Tables S1–S2.

## Notes

### Competing Interest Statement

The authors have declared no competing interest.

### Author Declarations

The Research Ethics Committee of the Hospital District of Northern Savo approved the study protocol in 2006 (Statement 69/2006). The parents or caregivers of the children gave their written informed consent, and the children provided their assent to participation. The PANIC study has been carried out in accordance with the principles of the Declaration of Helsinki as revised in 2008.

